# The efficacy and safety of argatroban with clopidogrel versus aspirin with clopidogrel for acute minor ischemic stroke (ACAP): study protocol for a multicenter, randomized controlled trial

**DOI:** 10.64898/2026.03.30.26349790

**Authors:** Huiting Zhang, Xiaotang Ma, Yuanliu Xiao, Geng Liao, Ning Kong, Tao Qin, Muqing Huang, Zhao Yin, Wenbin Chen, Jiayuan Wu, Junding Xian, Jiawu Fu, Feihong Xie, Congli Jin, Zhinmin Liao, Wei Liang, Lifeng Lin, Wenchuan Xian, Thanh N Nguyen, Duolao Wang, Wangtao Zhong, the ACAP investigators

## Abstract

**Background:** Previous studies have shown the benefit of dual antiplatelet therapy (DAPT) for acute minor ischemic stroke. Argatroban, is a thrombin inhibitor and is primarily used in patients with acute ischemic stroke experiencing early neurological deterioration. There is no study about the benefit of antiplatelet plus anticoagulant in this population. We aim to study the difference between the combination of argatroban and clopidogrel and DAPT in the outcomes of patients with acute minor ischemic stroke (AMIS, NIHSS ≤5) presenting within 72 hours after onset.

**Methods:** Argatroban combined with clopidogrel versus aspirin combined with clopidogrel in Stroke (ACAP study) is an investigator-initiated, multicenter, prospective, randomized, open-label trial with blinded endpoint evaluation conducted at four centers in China. This trial will randomize 464 eligible patients with minor ischemic stroke of NIHSS ≤ 5 (232 in each arm) within 72 hours of the last known well to receive intravenous argatroban with clopidogrel (treatment group) or aspirin plus clopidogrel (control group). The primary outcome is the proportion of patients achieving excellent outcome, defined as a score of 0-1 on the modified Rankin scale, at 90 days.

**Conclusions:** The ACAP trial will provide important data on the role of intravenous argatroban in patients with acute minor ischemic stroke presenting within 72 hours of last known well.

**Registration:** URL: https://www.chictr.org.cn; Unique identifier: ChiCTR2400080890; Date of registration: Feb 17, 2024.

## Introduction and rationale

Acute minor ischemic stroke (AMIS), characterized by a National Institutes of Health Stroke Scale (NIHSS) score less than or equal to 5, constitute approximately half of patients with acute ischemic stroke.^1–2^ AMIS is a significant cerebrovascular disease due to its high rate of early neurological deterioration (END) within 48-72 hours after AMIS onset, and a 90-day recurrent ischemic stroke risk ranging from 10% to 20%.^3–4^ Despite the established efficacy of reperfusion and thrombolytic therapies for acute ischemic stroke,^5^ constrained therapeutic windows and limited access to endovascular care^6^ preclude a subset of patients from receiving these interventions. Dual antiplatelet therapy (DAPT) is recommended for patients with AMIS,^7–8^ while the stroke recurrence rate remains relatively high (8.8-10.1%).^9^ Based on the natural history, it is imperative to explore new antithrombotic strategies for AMIS.

Argatroban, a direct-acting thrombin inhibitor, neutralizes both unbound and clot-associated thrombin. It is extensively employed in acute ischemic stroke therapy in Asia, notably China and Japan.^10^ Observational studies demonstrated that argatroban combined with recombinant tissue plasminogen activator (rtPA) improved neurological outcomes (mRS 0-1) at 90 days in patients with acute ischemic stroke, without increasing the risk of intracranial hemorrhage,^11^ whereas one randomized trial found that argatroban combined with intravenous thrombolysis did not improve clinical outcomes but instead increased mortality.^12^ Studies conducted in China and Japan have shown that argatroban combined with antiplatelet therapy reduced the incidence of END in patients with acute ischemic stroke.^9, 13^ A randomized trial showed that in patients experiencing END within 48 hours of symptom onset, argatroban plus antiplatelet therapy improved neurological deficits (mRS 0-3) at 90 days^10^ and reduced END in acute minor-to-moderate ischemic stroke due to large artery atherosclerosis and branch atherosclerosis disease with mild stroke.^14–15^ These results suggest that argatroban combined with antiplatelet therapy might be an effective strategy for patients with ischemic stroke. Currently, evidence regarding the efficacy and safety of argatroban combined with antiplatelet therapy in patients with AMIS within 72 hours of onset remains limited.

Hence, we designed a multicenter, open-label, blinded end point randomized trial of argatroban combined with clopidogrel versus aspirin combined with clopidogrel for acute ischemic stroke (ACAP), to test the hypothesis that argatroban combined with clopidogrel will improve clinical outcome in patients with AMIS presenting within 72 hours of last known well, and will not increase the incidence of symptomatic intracranial hemorrhage or mortality.

## Methods

### Study design

The ACAP study is an investigator-initiated, multicenter, prospective, randomized, open-label trial with blinded evaluation of outcomes (PROBE design), aiming to evaluate the superiority of intravenous argatroban plus clopidogrel (treatment group), compared to aspirin plus clopidogrel (control group), to increase the 90-day functional independence in AMIS patients who present within 72 hours of last known well (**Figure 1**). The trial is designed and conducted according to the Declaration of Helsinki and has been registered at https://www.chictr.org.cn, (identifier ChiCTR2400080890). The protocol (Version 3.0) was approved by the ethics committee of the Affiliated Hospital of Guangdong Medical University (Human Investigation Committee PJKT2023-101), and all participating hospitals before enrollment. The trial diagram and visit plan and the study flow chart are shown in **Figure 2 and Figure 3**. The enrollment, intervention, and evaluation schedule are shown in **Table 1**. The checklist of Standard Protocol Items: Recommendations for Interventional Trials (SPIRIT) is provided in Additional file 1.^16^

**Figure 1.**
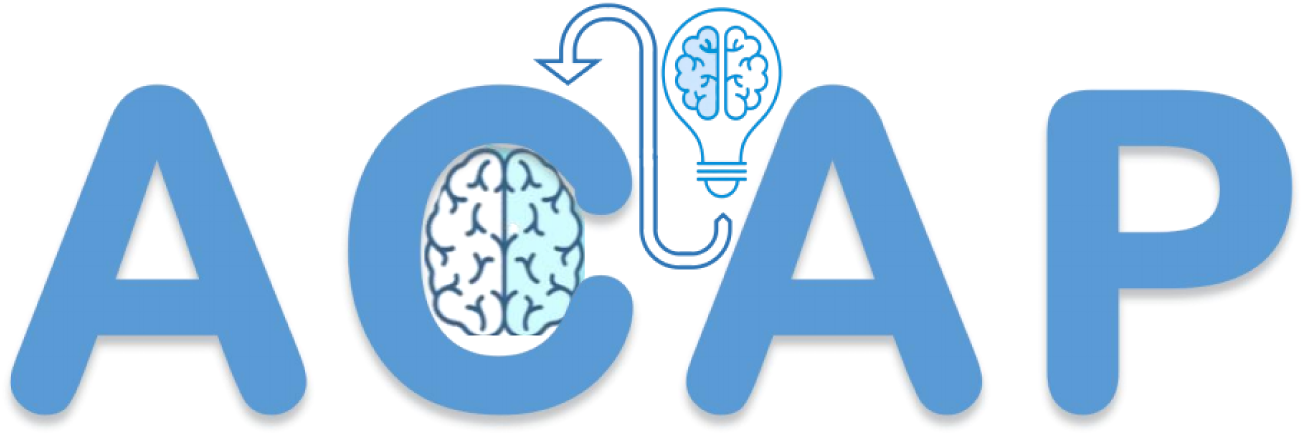
Trial logo.

**Figure 2.**
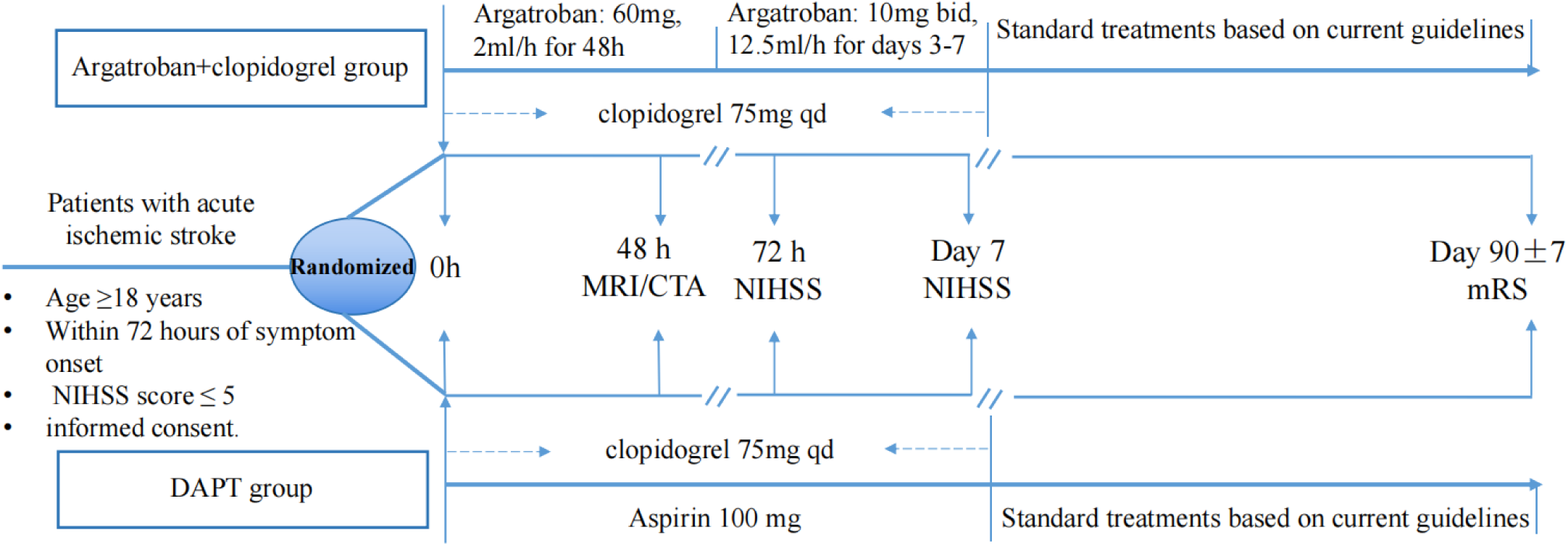
The trial diagram and visit plan.

**Figure 3.**
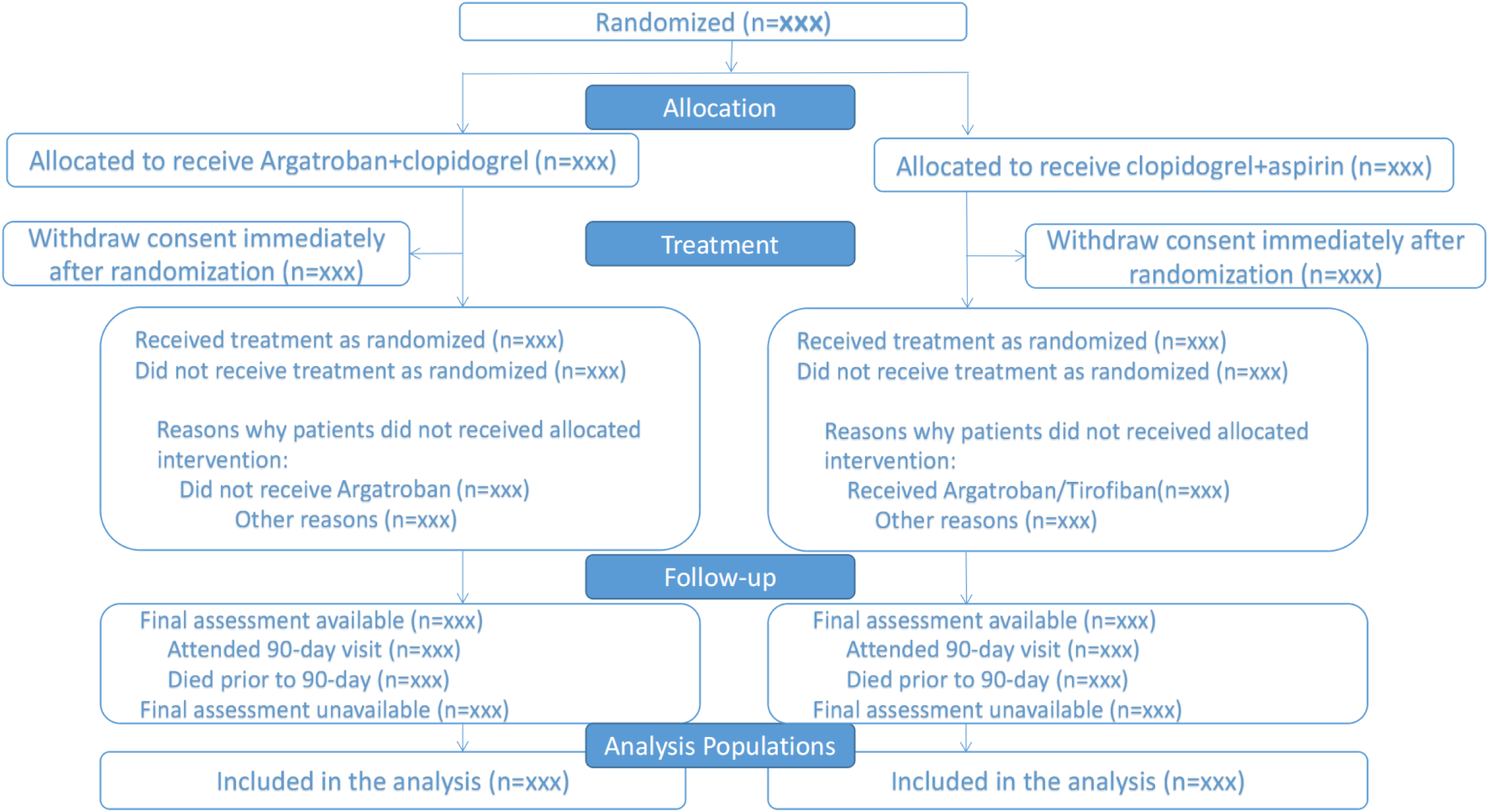
Study flow chart.

**Table 1.**
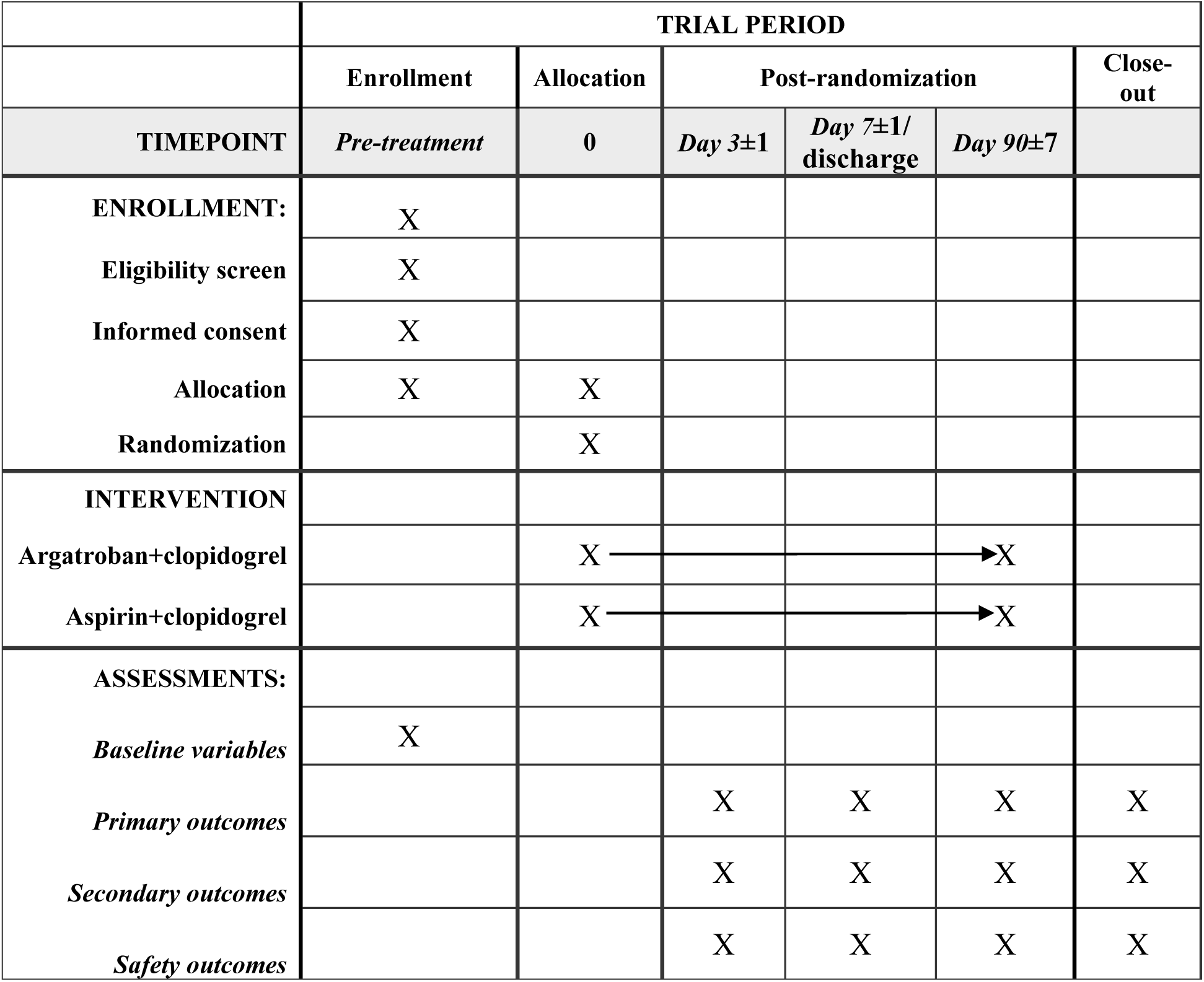
Participant timeline: Schedule of enrollment, interventions, and assessmentss.

## Recruitment

Eligible subjects will be consecutively enrolled across 4 Chinese stroke centers. To ensure the high-quality of the trial, protocol training was completed at all sites prior to patient enrollment.

### Participant population

#### Inclusion criteria include

(1) Age ≥18 years;
(2) Acute ischemic stroke patient with time of onset ≤ 72 hours and NIHSS score ≤ 5;
(3) Did not receive intravenous thrombolysis and/or mechanical thrombectomy;
(4) No significant pre-stroke functional disability: for age <80 years, pre-stroke mRS ≤2; for age ≥80 years, pre-stroke mRS ≤1;
(5) The patient or their legal representative provides written informed consent.

#### Exclusion criteria include

(1) History of atrial fibrillation;
(2) Coma;
(3) Intracranial hemorrhage on baseline CT or MRI;
(4) Already received intravenous thrombolytic after stroke;
(5) Known pregnancy, or breastfeeding, or serum beta human chorionic gonadotropin test is positive on admission;
(6) Current participation in another clinical trial;
(7) Patient with a preexisting neurological or psychiatric disease that would confound the outcome assessments;
(8) Mass effect or intracranial neoplasm on baseline CT or MRI (except small meningioma);
(9) Intracranial arteriovenous malformation or >3 mm unruptured aneurysm on baseline CT or MR angiography;
(10) Any terminal disease with a life expectancy less than half a year;
(11) Severe liver disease such as acute hepatitis, chronic active hepatitis, cirrhosis;
(12) Contraindications to argatroban, clopidogrel, or aspirin, including allergy, severe renal or hepatic insufficiency;
(13) History of thrombocytopenia or platelet count <100 x 10⁹/L, history of coagulopathy or systemic bleeding;
(14) Current use of anticoagulants or requirement for anticoagulant therapy during the study period;
(15) Gastrointestinal bleeding, genitourinary bleeding, or major surgery within the last 3 months;
(16) Individuals who are unable to cooperate or unlikely to be available for follow-up at 90 days

#### Randomization

The randomization list will be prepared by the independent statistical center using permuted-block randomization stratified by NIHSS scores (0-1, 2-3, 4-5). SAS 8.4 will be used to generate the randomization list. Sequentially numbered, opaque sealed envelopes containing computer-generated random assignment numbers and group allocation (control or treatment group) were prepared. Eligible enrolled patients were then assigned in a 1:1 ratio to the argatroban plus clopidogrel group or aspirin plus clopidogrel group according to the random number in the envelope. Both the patient and physician will be aware of the treatment assignment, and the assignment personnel will have access to treatment assignment. Other research team members such as enrollment personnel, data managers, and statisticians will be blinded about the treatment allocation. The follow-up NIHSS assessments will be performed by the treating physician. Trained investigators will evaluate the mRS score at 90 days using a structured questionnaire in a blinded fashion.^17^

## Assignment of interventions: blinding and unblinding

Outcome assessors and data analysts will be masked to group allocation. Participants will be instructed not to disclose their assignment to assessors. Emergency unblinding was not required, as this study is an open-label RCT with blind endpoint assessment.

### Treatments

#### Argatroban plus clopidogrel group

Patients in the argatroban plus clopidogrel group will receive treatment as follows: 75 mg clopidogrel administered orally on the first day and continued daily for 90 days. Argatroban (10mg/2mL) is supplied by Jiangsu Desano Pharmaceuticals Co., LTD. For days 1 to 2, 60 mg of argatroban dissolved in 40 mL of Normal Saline (NS) is administered to the patient via intravenous infusion pump at a rate of 2 mL/h. For days 3 to 7, 10 mg of argatroban dissolved in 40 mL of NS is administered to the patient via intravenous infusion pump at 12.5 mL/h or in NS 250 mL via intravenous infusion at a rate of 20 drops per minute, twice daily.^10^ The regimen is empirically optimized from prior trial data (ARTSS-2, ARGIS-1)^18–19^ to maximize early thrombin inhibition while minimizing bleeding.

Argatroban infusion rates will be adjusted to achieve a target activated partial thromboplastin time (APTT) of 1.75 × baseline (±10%).^11^ Patient adherence to the continuous intravenous argatroban infusion protocol will be rigorously monitored. This includes continuous electronic recording of infusion pump data and verification against prescribed dosing parameters. The nursing staff will perform regular checks every two hours.

#### Aspirin plus clopidogrel group

Patients assigned to the aspirin plus clopidogrel group will receive treatments as follows: 100 mg aspirin and 75 mg clopidogrel are administered orally on the first day, and continued daily for 7 days, and 75 mg clopidogrel administered orally from days 8 to 90.

All enrolled patients in both groups will receive treatment as per the current American or Chinese stroke guidelines.^20–21^ Patients receiving argatroban undergo a head CT scan and coagulation function tests with any deterioration of NIHSS.

## Relevant concomitant care and interventions during the trial

The use of other anticoagulant/antiplatelet agents, such as tirofiban, indobufen, sulodexide, dabigatran etexilate, rivaroxaban and heparin, is prohibited during the trial period.

### Efficacy end-points

The primary endpoint is excellent outcome, defined as a modified Rankin scale (mRS) score of 0 to 1 at Day 90 (±7) after randomization.

The secondary endpoints are:

(1) Disability level measured by the mRS at 90±7 days;
(2) Functional independence defined as mRS 0 to 2 at 90±7 days;
(3) Independent ambulation defined as mRS 0 to 3 at 90±7 days;
(4) END defined as an increase in NIHSS score≥4 points within 72 hours of randomization;^22^
(5) Recurrent stroke (ischemic/hemorrhagic stroke, TIA) or other vascular event within 90 days.
(6) Median NIHSS score at 7 days or discharge if earlier;
(7) Health-related quality of life [European Quality Five-Dimension Five-Level (EQ-5D-5L) scale score] at 90±7 days.

### Safety end-points

The primary safety outcome is symptomatic intracranial hemorrhage (SICH) within 7 days after randomization. SICH will be adjudicated by an independent Imaging Core Laboratory according to the modified Heidelberg Bleeding Classification, defined as any hemorrhage with neurological worsening with an increase in NIHSS score of ≥4 points compared to baseline or the lowest value in the first 7 days or any hemorrhage resulting in death, as specified in the trial protocol.^23–24^ SICH will be confirmed on independent reading by blinded central study radiologists.^24^

### The secondary safety outcomes are as follows

(1) All-cause mortality at 90±7 days;
(2) Serious adverse events;
(3) Bleeding Academic Research Consortium (BARC) types 2, 3, and 5 bleeding event within 7 days.^25^
(4) Asymptomatic intracerebral hemorrhage (aICH, defined as any intracerebral hemorrhage that did not meet the criteria for SICH) event within 90±7 days.^26^

## Data collection plan

Prior to study commencement, to ensure the research quality, we will train recruiting physicians and assessors, and implement the following measures: The recruiting physicians will achieve proficiency in the experimental protocol and managing potential adverse events during therapy. Assessors will undergo training in the standardized acquisition and analysis of outcome measures.

## Plans to promote participant retention and complete follow-up

Follow-up assessments will be conducted on day 3, day 7 (or at discharge, whichever occurs first), and day 90 after enrollment. Participants will be contacted via telephone and asked to either return to the hospital or to complete the items within the assessment scales.

### Data and safety monitoring board

The independent data and safety monitoring board (DSMB) is composed of 3 specialists in stroke and biostatistics, all of whom do not participate in the ACAP trial and are external to the study sponsors. Strict measures will be implemented to safeguard the privacy rights of each participant. Initial data and outcome indicators will be securely stored in a highly protected database, and anonymization procedures will be rigorously followed to ensure the safety and confidentiality of all participants. The DSMB will meet once a year to monitor the progress of the trial. The DSMB will review the frequency (percentage) of serious adverse events and provide recommendations on whether to suspend, continue, or stop the trial. The DSMB convened on January 5, 2025, to conduct a scheduled interim review of the clinical trial. Following comprehensive evaluation of accumulated safety data, efficacy endpoints, and protocol adherence, the DSMB recommended continuation of the trial.

## Composition of the coordinating center and trial steering committee

The Department of Neurology at Affiliated Hospital of Guangdong Medical University will serve as the trial coordinating center. The steering committee, co-chaired by WTZ and HTZ, is responsible for protocol ratification, providing methodological oversight, and monitoring trial progression. HTZ, GL, YLX, and MQH will manage regulatory affairs, including securing ethics committee approval and coordinating audit compliance. HTZ, GL, YLX, WCX, ZY, MQH, JWF, FHX, CLJ, ZML, WL, LFL and WBC are assigned to participant recruitment, screening, and enrollment, with HTZ additionally supervising treatment administration. Data acquisition and statistical analysis are entrusted to TNN and DLW. The project management team convenes every two weeks to monitor the advancement of the trial. The steering committee meets biweekly to deliver strategic oversight to the investigative team, ensuring protocol adherence and uninterrupted trial implementation.

## Adverse event reporting and harms

The research team will strictly comply with all applicable regulations stipulated by the medical ethics committee to ensure the paramount safety and ethical protection of study participants. Any adverse events (e.g., gingival bleeding, gastrointestinal reactions, hemorrhagic transformation) occurring during the study period, whether related to the study intervention or not, will be promptly documented and addressed. In the event of an adverse reaction, the investigational drugs will be immediately discontinued, and the affected participant will receive appropriate medical treatment.

## Plans for protocol amendments

The principal investigator (PI) will notify the coordinating centers and ensure the prompt dissemination of the revised protocol. The clinical trial registry will implement the corresponding updates to accurately reflect the amended procedures.

### Sample size estimates

The sample size was calculated based on the estimated treatment effects based on a binary assessment of excellent functional outcomes (mRS score 0-1) at 90 days. A randomized clinical trial showed that argatroban contributed to a 19.2% absolute increase in the proportion of excellent outcome at 90 days compared with DAPT in AMIS patients with branch atherosclerosis disease (median NIHSS was 2).^15^ In acute ischemic stroke patients who experienced END (median NIHSS was 8), argatroban had a 6.9% absolute increase in the proportion of excellent outcome at 90 days compared with DAPT.^10^ Argatroban plus r-tPA resulted in a 9% improvement in the excellent outcome at 90 days compared to r-tPA alone in acute ischemic stroke patients (median NIHSS was 8).^11^ Therefore, in this study, we hypothesized that argatroban would result in a 9% absolute increase in the proportion of patients (NIHSS≤5) achieving excellent outcome at 3 months (mRS score 0-1) compared with DAPT. The difference in outcome between the two treatment groups was estimated with data from an acute stroke registry showing that patients with minor ischemic stroke who received DAPT treatment had a 3-months mRS 0-1 of 82.9%.^27^ With power set at 80% and an α level at 0.05, the estimated sample size was 418 patients. Adding 10% loss to follow-up and adjusting for a single interim analysis for efficacy, the sample size estimate was increased to 464 patients (232 in each treatment group). This estimate was conducted using PASS software (NCSS, LLC. Kaysville, Utah, USA) version 15.0.

## Trial results dissemination plan

The investigators commit to the timely communication of clinical trial results to participants, healthcare professionals, and the public through the clinical trial registries. Results will be submitted to Chinese Clinical Trial Registry, https://www.chictr.org.cn) within 12 months of trial completion.

### Statistical analysis

Primary trial analyses of efficacy outcomes will be on the intention to treat population and additional analyses will be performed on the per protocol population. For the analyses of the primary outcome and secondary binary outcomes, the modified Poisson regression with robust error estimation will be used.^28^ An ordinal outcome such as mRS (0-6) at 90 days will be analyzed using the generalized odds ratio (GenOR) approach. Continuous secondary outcomes such as the EQ-5D-5L scale score will be analyzed using linear regression model or the win ratio method depending on their distributions.^29^ In addition, covariate adjusted analyses of the primary and secondary outcomes will be performed adjusting for baseline NIHSS score, age, baseline ASPECTS (Alberta Stroke Program Early Computed Tomography Score), occlusion site, and time from last known well to randomization. Model-based covariate adjusted analyses will be performed, and if a covariate adjusted model does not converge or is unavailable, inverse probability treatment weighting method will be employed. Unadjusted and adjusted risk ratios, mean differences/win ratios, and generalized odds ratios will be calculated with their corresponding 95% confidence intervals. The missing values in primary and secondary outcomes will not be imputed, but sensitivity analyses based on different hypotheses about the missingness patterns of the primary outcome will be performed to assess the robustness of the primary outcome analysis results. Safety outcomes in both groups will be reported as frequency counts and percentages. All analyses will be detailed in the statistical analysis plan which will be finalized before unblinding the study data. The significance level for all analyses is a two-sided α of 0.05. Statistical analyses will be conducted on the SAS 9.4 system with Windows SAS (version 9.4) or R (version 4.3.1).

In predefined subgroup analyses, we explored differences in outcomes for the following parameters: NIHSS score (<3 or ≥3), time from symptom onset of event to randomization (≤24 hours or >24 hours), age (<65 or ≥65 years old), sex (male or female), Trial of ORG 10172 in Acute Stroke Treatment subtypes, ASPECTS, prior stroke, prior antiplatelet, occlusion site (anterior circulation, posterior circulation), relevant artery diseases with moderate-to-severe stenosis (luminal narrowing ≥50%), smoking, diabetes, hypertension, baseline NIHSS scores.

### Trial status

The trial was registered on Feb 17, 2024 and is ongoing. At the time of submission of this article, 464 patients were randomized. Enrollment is expected to end in August 2026, while follow-up for the final patient is targeted to finish in November 2026.

## Discussion

The ACAP randomized trial will provide important data on the efficacy and safety of argatroban in AMIS, and will explore whether extending the time window for argatroban therapy to within 72 hours after onset of symptoms would be beneficial for AMIS patients.

Despite receiving guideline-recommended DAPT, some AMIS patients still develop vascular events or recurrent stroke. The synergistic combination of the direct thrombin inhibitor argatroban with an antiplatelet drug may enhance microvascular perfusion and improve patient prognosis. The study investigators considered it ethical for both groups to discontinue DAPT from days 8 to 21 and extending the therapeutic window for argatroban to 72 hours. First, this trial was designed and registered in February 2024 based on results of the argatroban plus rtPA for acute ischemic stroke (ARAIS and ARTSS-2) studies,^11, 18^ demonstrating the efficacy or safety of argatroban. It was also aligned with Chinese expert consensus on argatroban for acute ischemic stroke. The enrolled participants of our trial were diagnosed with AMIS within 72 hours of onset, while current guidelines recommend prompt initiation of DAPT within 24 hours and last for 21 days based on the results of the CHANCE and POINT trials.^7–8^ A Korean investigation revealed that from 2008-2022, approximately 33% to 78% of AMIS patients presenting more than 24 hours after symptom onset still received DAPT. The study found that these patients had a reduced composite risk of stroke, myocardial infarction, and all-cause mortality within 3-month.^30^ This practice was supported by a subsequent randomized trial of 6100 patients which showed that the optimal initiation window for DAPT might be within 72 hours of symptom onset.^31–32^ A systematic review and meta-analysis indicated that initiating DAPT within 7 days may still confer clinical advantage for individuals with acute ischemic stroke.^33^ Second, argatroban significantly increased the proportion of mRS 0-2 in patients with mild-to-moderate ischemic stroke with large artery atherosclerosis within 72 hours at 90 days compared with antiplatelet therapy (85.3% vs 74.5%, p=0.04).^14^ No statistical differences were observed in hemorrhagic transformation of infarction and systemic hemorrhage between the two groups, suggesting the relatively safety of argatroban treatment within 72 hours of stroke onset. A prospective study showed that the 90-day risk of composite vascular events including ischemic stroke recurrence, transient ischemia attack, symptomatic intracerebral haemorrhage, myocardial infarction or angina attacks and vascular death in patients with AMIS who received a shorter duration of DAPT (less than 10 days) was 10.2%.^34^ The INSPIRE study showed that AMIS patients receiving 21 days of DAPT therapy had a 7.5% risk of composite vascular events including stroke, myocardial infarction, or death from cardiovascular causes within 90 days.^32^ Treatment with DAPT yielded a decrease in ischemic stroke incidence within the first 14 days. Beyond day 10, the incidence of hemorrhagic events induced by DAPT surpassed the number of strokes prevented by these regimen.^35^ Therefore, a short duration of DAPT is not likely to substantially disadvantage patients in ACAP. Last, The INSPIRE trial also demonstrated that in patients with AMIS within 72 hours of onset, standard DAPT therapy reduced the 90-day stroke recurrence compared to aspirin monotherapy (HR, 0.79, 95% confidence interval, 0.66-0.94; P=0.008). However, the risk of moderate-to-severe bleeding within 90 days was significantly higher in the DAPT than aspirin group (HR, 2.08; 95% CI, 1.07-4.04; P=0.03).^32^ These findings indicate that though DAPT is superior to aspirin monotherapy in AMIS patients within 72 hours of onset, the modest increase in the risk of moderate-to-severe bleeding should be considered. Given that the trial regimen included argatroban combined with clopidogrel for the first 7 days, extending DAPT to 21 days might potentially confer a greater bleeding risk. Therefore, DAPT was discontinued from days 8 to 21 and the Ethics Committee approved the ACAP trial.

This trial has several limitations. First, because argatroban dosing requires titration guided by APTT monitoring, the design of our trial is open label. Therefore, we opted for the PROBE design to reduce bias by blinding endpoint assessors for the 90-day clinical endpoints. Second, this study excluded patients with cardioembolic ischemic stroke, leading to a study cohort predominantly consisting of atherosclerotic-type patients. Third, argatroban requires a continuous intravenous administration, which may be less convenient to patients as compared to oral aspirin. In addition, the generalizability of our findings is limited due to individual variations in ischemic stroke, the geographic constraints of our participants within Western Guangdong, China. Therefore, further validation is required in national and international populations.

## Conclusions

In summary, ACAP is designed to determine the role of argatroban in addition to clopidogrel within 72 hours of symptom onset in patients with acute minor ischemic stroke. This trial will contribute to the evidence for the adjunct use of argatroban plus clopidogrel compared with aspirin plus clopidogrel within a 72-hour time window.

## Ethics approval and consent to participate

This study was approved by the ethics committee of the Affiliated Hospital of Guangdong Medical University and all participating centers. (Human Investigation Committee PJKT2023-101). Written informed consent to participate will be obtained from all participants, and the informed consent is provided in the supplementary materials.

## Availability of data and materials

Data will be accessible to researchers affiliated with academic or non-profit institutions, regulatory authorities upon legitimate request and other investigators whose proposed use aligns with the original study objectives. The data that support the findings of this study are available from the corresponding author upon reasonable request.

## Authors’ contributions

The project design was carried out by HTZ, XTM and WTZ, while the protocol modification was executed by DLW, TNN and HTZ. Feasibility analysis was conducted by HTZ and DLW, with recruitment and evaluation of trial participants jointly managed by HTZ, YLX, GL, NK, TQ, MQH, ZY, WBC, JYW, JDX, JWF, FHX, CLJ, ZML, WL, LFL, WCX, and WTZ. The intervention measures will be supervised and completed by HTZ. Statistical analysis will be performed by XTM and DLW, while trial execution monitoring will be the responsibility of WTZ. HTZ and XTM wrote the manuscript, which was revised by DLW, TNN and WTZ. All authors read and approved the final manuscript.

## Disclosure

None.

## Abbreviations

DAPT: Dual antiplatelet therapy
AMIS: Acute minor ischemic stroke
NIHSS: National Institutes of Health Stroke Scale
END: Early neurological deterioration
APTT: activated partial thromboplastin time
ASPECTS: Alberta Stroke Program Early Computed Tomography Score
BARC: Bleeding Academic Research Consortium
EQ-5D-5L: European Quality Five-Dimension Five-Level

